# Neurophysiological consequences of synapse loss in progressive supranuclear palsy

**DOI:** 10.1101/2022.06.22.22276697

**Authors:** Natalie E. Adams, Amirhossein Jafarian, Alistair Perry, Matthew A. Rouse, Alexander D. Shaw, Alexander G. Murley, Thomas E. Cope, W. Richard Bevan-Jones, Luca Passamonti, Duncan Street, Negin Holland, David Nesbitt, Laura E. Hughes, Karl J Friston, James B. Rowe

## Abstract

Synaptic loss occurs early in many neurodegenerative diseases and contributes to cognitive impairment even in the absence of gross atrophy. Currently, for human disease there are few formal models to explain how cortical networks underlying cognition are affected by synaptic loss. We advocate that biophysical models of neurophysiology offer both a bridge from clinical to preclinical models of pathology, and quantitative assays for experimental medicine. Such biophysical models can also disclose hidden neuronal dynamics generating neurophysiological observations like electro- and magneto-encephalography (MEG). Here, we augment a biophysically informed mesoscale model of human cortical function by inclusion of synaptic density estimates as captured by [^11^C]UCB-J positron emission tomography, and provide insights into how regional synapse loss affects neurophysiology. We use the primary tauopathy of progressive supranuclear palsy (Richardson’s syndrome) as an exemplar condition, with high clinicopathological correlations. Progressive supranuclear palsy causes a marked change in cortical neurophysiology in the presence of mild atrophy and is associated with a decline in cognitive functions associated with the frontal lobe. Using (parametric empirical) Bayesian inversion of a conductance-based canonical microcircuit model of MEG data, we show that the inclusion of regional synaptic density—as a subject-specific prior on laminar specific neuronal populations—markedly increases model evidence. Specifically, model comparison suggests that a reduction in synaptic density in inferior frontal cortex affects superficial and granular layer glutamatergic excitation. This predicted individual differences in behaviour, demonstrating the link between synaptic loss, neurophysiology, and cognitive deficits. The method we demonstrate is not restricted to progressive supranuclear palsy or the effects of synaptic loss: such pathology-enriched dynamic causal models can be used to assess the mechanisms of other neurological disorders, with diverse non-invasive measures of pathology, and is suitable to test the effects of experimental pharmacology.

## Introduction

Human neurodegenerative diseases are heterogeneous in their symptoms, progression and molecular biology but they all call for mechanistic explanations of pathophysiology underlying cognitive impairment^1–4^. This may be met by biophysically informed models of brain-network dynamics that integrate patient-specific measures of neuropathology. We propose that by embedding neuropathological information in individualised disease models, one could establish bridges between clinical and preclinical models of disease, facilitate experimental medicine, and inform precision medicine. We therefore sought to enrich biophysically informed generative models of cortical neurophysiology, inverted from magnetoencephalography, with markers of neuropathological severity from positron emission tomography.

We focus on synapse loss as the neuropathology, which is common across many neurodegenerative diseases, and closely related to the severity of dementia^5–12^. This kind of synapse loss is a consequence of protein misfolding, aggregation and inflammation in multiple disorders, and begins before cell death^13^. Post-mortem studies have identified cell- and region-specific changes in synaptic density^14–16^. Quantification of region-specific synaptic density is now possible *in vivo* with PET, using ligands for the presynaptic vesicle protein 2A (SV2A PET)^12,17,18^. However, less is known about the impact of this synaptic loss on the neurophysiological function of local cortical networks^7^.

To characterise the relationship between synaptic loss and cortical neurophysiology, we use the primary tauopathy of Progressive Supranuclear palsy (PSP) as an exemplar condition. Within the group of frontotemporal lobar degeneration pathologies, PSP has very high clinicopathological correlation. Over and above the motor impairments of PSP, it is associated with marked decline in cognitive function and physiological responses, especially cognitive functions associated with the frontal lobe^19–21^. These frontal physiological and cognitive changes occur in Richardson’s syndrome as well as the PSP-Frontal phenotype, despite only mild cortical atrophy^22^. The discrepancy between severe functional deficits and mild atrophy has been proposed to result from changes in synaptic density and loss of major neurotransmitter systems in the frontal lobe^22–27^. PSP synaptic loss is severe in multiple cortical regions at *post mortem* and *in vivo*^10,28^, making the disorder ideally suited to demonstrate the relationship between synaptic loss and cortical function.

We had three principal aims. First, to develop a method for pathology-enriched dynamic causal models (DCM), combining magnetoencephalography with PET data. Using this method, we could test for a relationship between synaptic density and inferred synaptic efficacy within the generators of magnetoencephalography signals. Second, we hoped to identify the subject-, layer- and cell-specific parameters that are most sensitive to changes in synaptic density. We focus on the synaptic loss and neurophysiological function of the frontal lobe (specifically inferior frontal gyrus, IFG) because of the cognitive profile of PSP. The third aim was to test the hypothesis that the neuronal parameter estimates would be correlated with cognitive function, endowing them with construct validity.

In pursuing these aims, we paid careful attention to the validity of the modelling and data analysis: face validity was established in terms of the predictive accuracy (i.e., the model could reproduce realistic neurophysiological responses). Construct validity was addressed through the use of independent measures of psychopathology and pathophysiology. Finally, predictive validity was addressed with a quantitative analysis of reliability using the Intraclass Correlation Coefficient and a split half procedure (i.e., odd and even trials).

## Materials and Methods

### Participants

Eleven people with probable PSP Richardson’s syndrome^29^ underwent structural MRI, [^11^C]UCB-J PET and magnetoencephalography. Whereas prominent presenting features can be cognitive and behavioural (e.g. in PSP-Frontal phenotype), all had progressed to Richardson’s syndrome by the time of the study. Participants were recruited from the Cambridge Centre for Parkinson-plus and gave written informed consent in accordance with the Declaration of Helsinki (1991). Their clinical and cognitive assessment included the Mini Mental State Examination (MMSE), revised Addenbrookes Cognitive Examination (ACE-R,) Cambridge Behavioural Inventory (CBI-R), Hayling sentence completion test, INECO Frontal Screening (IFS), Progressive Supranuclear Palsy Rating Scale (PSPRS), Frontal Assessment Battery (FAB), and Graded Naming Test. Demographic and clinical data of participants are summarised in Supplementary Table 1.

### Neuroimaging data acquisition

During magnetoencephalography, participants were exposed to a roving auditory oddball stimulus train, as described in Adams *et al*.^19^. Magneto/electrophysiological data were recorded at 1000 Hz using a 306-channel Vectorview acquisition magnetoencephalography system (Elekta Neuromag, Helsinki) located in an Elekta Neuromag magnetically-shielded room. Sensors are in triplets, as a pair of gradiometers and a magnetometer. Electrooculograms tracked eye movements vertically and horizontally, and 5 head-position indicator coils tracked the head position (500 Hz). EEG was simultaneously recorded using a 70 channel, MEG-compatible, EEG cap (Easycap GmbH). A 3D digitizer (Fastrak Polhemus Inc., Colchester, VA) recorded >100 scalp points, nasion and bilateral pre-auricular fiducial points.

For co-registration with the MEG data, T1-weighted structural MRI was collected in a 7T Siemens TERRA scanner (MP2RAGE sequence, TE=1.99ms, TR=4300ms, 0.75mm isotropic voxels). For one subject the scan was collected in a 3T Siemens PRIMSA scanner (with magnetization-prepared rapid gradient-echo (MPRAGE sequence sampling, echo time (TE) = 2.9ms, repetition time (TR) = 2000ms, 1.1mm isotropic voxels) at the Wolfson Brain Imaging Centre, University of Cambridge.

Participants underwent a [^11^C]UCB-J PET scan, on a GE SIGNA PET/MR (GE Healthcare, Waukesha, WI), with 90 minutes dynamic imaging following [^11^C]UCB-J injection, with attenuation correction including the use of a multi-subject atlas method^30^ and improvements to the brain MRI coil component. Full details of the post-processing are provided in Holland *et al*.^10^ In brief, the data were attenuation corrected and aligned to a simultaneous subject-specific T1-weigthed MRI, TE = 9.2 msec, TR = 3.6 msec, 1.0 mm isotropic voxels, 192 sagittal slices, in-plane voxel dimensions 0.55×0.55 mm (subsequently interpolated to 1.0×1.0 mm); slice thickness 1.0 mm).

Regions were specified using the Hammersmith Atlas. Regional time–activity curves were extracted following the application of geometric transfer matrix partial volume correction to each dynamic image. Regions of interest were multiplied by a binary grey matter mask (>50% on the SPM12 grey matter probability map smoothed to PET spatial resolution). The non-displaceable binding potential of [^11^C]UCB-J was estimated as the measure of synaptic density, using the simplified reference tissue model with the centrum semiovale as the reference region (corrected for CSF and grey matter partial volume). Only the right IFG data were carried over to this study.

### Data pre-processing

Magnetoencephalography data were acquired using a standard (roving) auditory mismatch negativity (MMN) paradigm. The data were MaxFiltered (v2.2, Elekta Neuromag) to remove external noise, correct for head motion and interpolate bad channels. Subsequent data processing was performed with the Statistical Parametric Mapping toolbox (SPM12 v7771, Wellcome Trust Centre for Neuroimaging, UK) FieldTrip (fieldtriptoolbox.org) and OSL (https://github.com/OHBA-analysis/osl-core) software in MATLAB (2019a, Mathworks, Natick, MA). Data were downsampled to 500Hz, band-pass filtered between 0.1-125 Hz and notched between 45-55Hz and 95-105Hz. Bad channels were removed using *osl_detect_artefacts*, before independent component analysis was used to remove eye-motion artefacts. Data were then epoched from −100 ms to 400 ms relative to stimulus onset. Further artefact rejection used thresholding of MEG channels to remove bad trials (*osl_detect_artefacts*). The deviant and standard trials—that constitute the roving MMN paradigm—were taken as the 1st and 6th trials of each stimulus train respectively, following a change in auditory tone.

Conventional source reconstruction was performed using the COH (i.e. coherence) method in SPM12, using subject specific structural images. Source data timeseries were obtained using a region of interest—with a radius of 7mm—for the reconstruction of regional responses. A single (representative) source was selected for subsequent analysis of between subject differences: namely, the right inferior frontal gyrus (RIFG), with the Montreal neurological institute template coordinate of [46, 20, 8]. This is the prefrontal source in the auditory hierarchy (of five sources) that generate the auditory evoked responses (and accompanying differences that constitute the MMN).

### Dynamic Causal Modelling

A conductance-based DCM with six neuronal populations or cell types was used, modified from the CMC-NMDA model as described in Adams *et al*.^31^. A list of the mean and variance of prior model parameters is provided in Supplementary Table 2. These parameters pertain to synaptic time constants and rate constants that parameterise the efficacy of intrinsic (i.e., within source) connections among the six populations, which are usually assigned to cortical lamina. The RIFG event-related field (ERF) timeseries for each participant was used for model inversion using standard (variational Laplace) procedures (with a maximum of 64 iterations of the inversion scheme).

The ensuing parameter estimates for each participant were entered into parametric empirical Bayes (PEB) analyses. These between subject analyses were used to test the hypothesis that one or more synaptic parameters could be explained by differences in (PET derived) synaptic density measures from the RIFG region of interest. The ensuing (PEB) models were created from 6 sets of synaptic parameters: superficial and deep AMPA, NMDA and GABA. These 2 × 3 = 6 groups form a model space of 63 models (as 2^6 – 1 = 63), where each model corresponds to a particular combination of synaptic parameters that could be influenced by synaptic density. The evidence (a.k.a., marginal likelihood) for each model was evaluated using the variational evidence lower bound (i.e., variational free energy). The resulting free energies were converted to probabilities over models via the softmax operator. In this work, we focused on the synaptic parameters mediating responses to both standard and deviant stimuli, where the differential responses (that underwrite the MMN) were modelled with parameters, mediating condition-specific changes in connectivity. This allowed us to use the amplitude in the MMN window as an independent marker of disease severity as follows.

A further PEB analysis was undertaken to assess the contributions of two independent measures; namely, (i) the cognitive deficits as measured by the Frontal Assessment Battery; chosen because of its clinical utility, sensitivity to the presence of PSP and association with frontal lobe pathology; and (ii) a simple evoked physiological response, quantified here as the single maximal deflection during the mismatch window (130–180ms, noting that the DCM inversion uses the full time series of standard and deviant tones, and not a singular parameter of the peak MMN response). These PEB analyses looked for influences on the above (six) sets of superficial and deep AMPA, NMDA and GABA connections.

Finally, the above analyses were re-run for odd and even trials separately, to create independent datasets and subsequent model inversion. The Intraclass Correlation Coefficient (ICC^32^) was used as a measure of the within-subject reliability, using the odd/even trials’ data.

### Data Availability

The MEG data pre-processing pipeline is available at https://github.com/-AlistairPerry/FTLDMEGMEM. The DCM used in this study was adapted from https://gitlab.com/tallie/edcm, as described in Adams *et al*.^31^ with prior model parameters altered according to Supplementary Table 2.

## Results

The observed ERFs and model ERF predictions (Fig. 1A) were highly correlated (Fig. 1B, mean Pearson’s correlation = 0.86 ±0.15), reflecting the face validity of this DCM.

**Figure 1.**
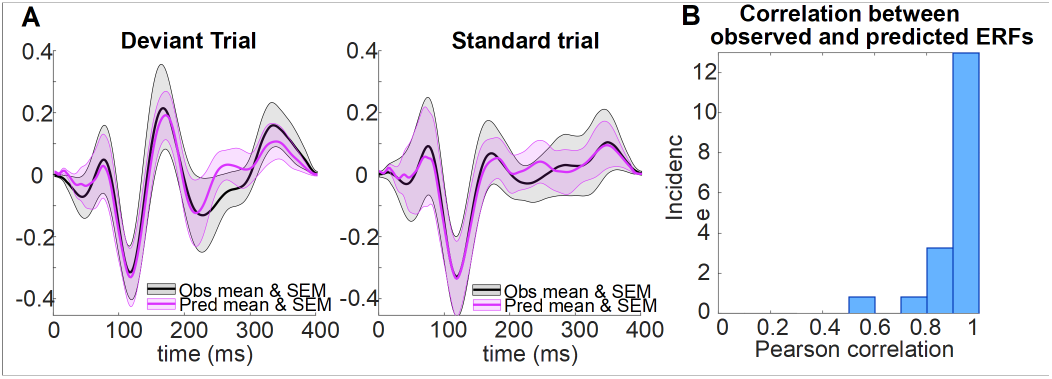
The accuracy of the generative model of the evoked response. A) ERFs for the standard and deviant trials shown as a mean with SEM over all subjects. Observed data is in black and data predicted by the canonical microcircuit model is in purple. B) The histogram illustrates the high correlations between observed and predicted ERFs for each participant.

To identify the relationship between each participant’s regional synaptic density and the DCM estimates of synaptic efficacy, PEB was used to search for the best mapping from subject specific synaptic density in RIFG to different combinations of synaptic parameters in the DCM of the same region. These between subject (PEB) models were compared using their free energy (Fig. 2A, with the model space described in the lower matrix). The model space covers all combinations of superficial and deep AMPA, NMDA and GABA connections. The winning model of all available data (Fig. 2A, *top*) was the model in which superficial AMPA and NMDA synaptic connections were sensitive to PET measures of synaptic density (posterior probability = 0.52), followed closely by a model that also included deep NMDA connections (posterior probability = 0.32).

**Figure 2.**
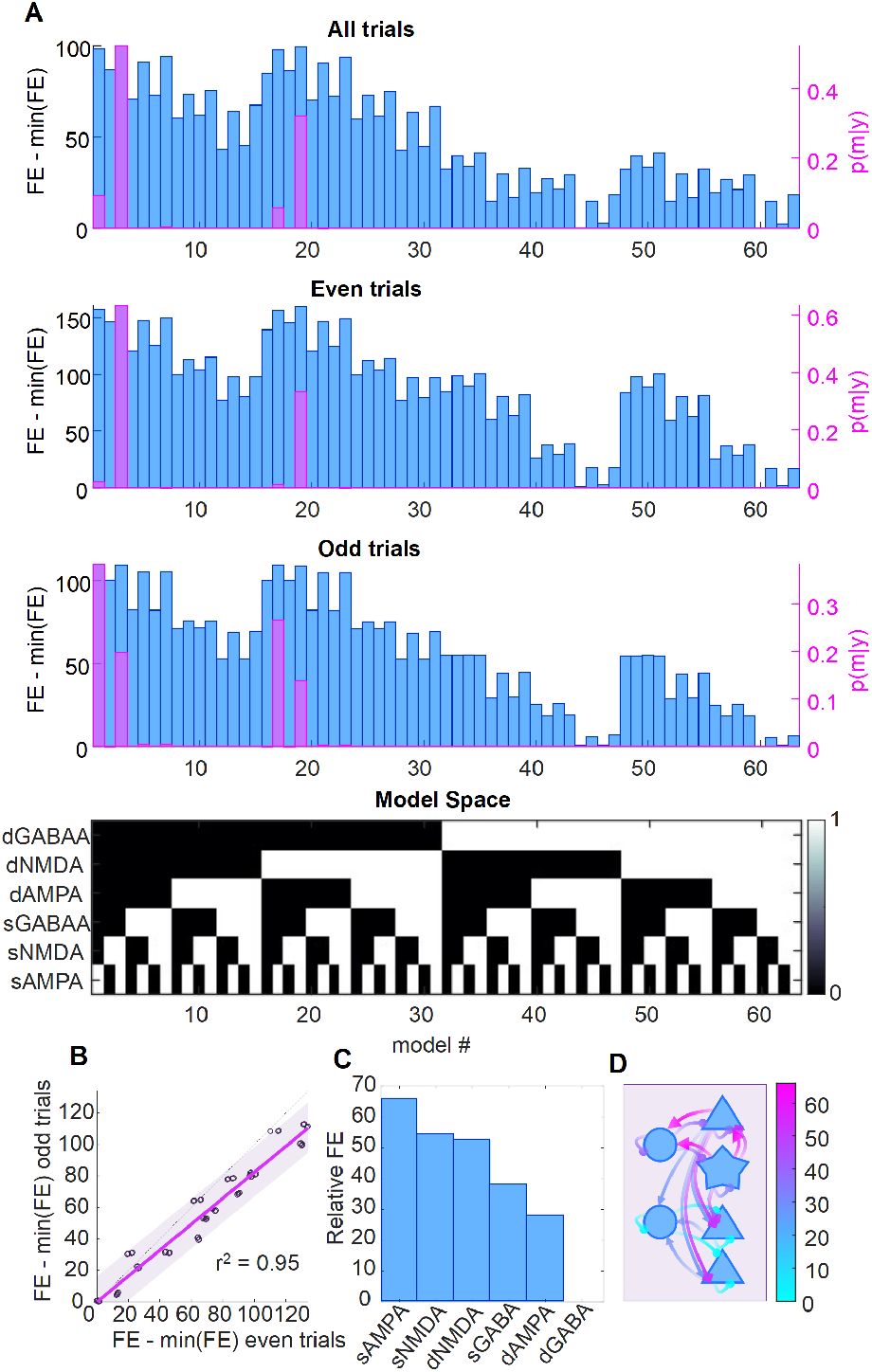
The reliability of the free energy estimation over independent data per subject and over subjects. A) The free energy (FE, blue bars) for all 63 models for all trials (top), even trials (middle) and odd trials (bottom). The righthand y-axis (and pink bars) shows the posterior probability of each model. The model space, aligned with the bar charts above, is shown as the black and white matrix below. B) A comparison of the free energy for the most likely model plotted separately for even trials and odd trials for each subject, indicating the high reliability of the free energy estimate of the bound on (log)-model evidence. C) The relative free energy for each connection group when considered in isolation. D) The relative free energy for each connection group, with arrows distinguishing synapse type (triangle = AMPA; circle = GABA; diamond = NMDA). The scale for the colormap refers to the range of relative free energies across the groups.

To assess the reliability of the PEB analyses, DCM was applied separately for odd and even trials. Results for ‘all’, ‘odd’ and ‘even’ trials are reported in Fig. 2A. The free energies of all models for ‘odd’ and ‘even’ conditions were highly correlated (r^2^ = 0.95, Fig. 2B). The overall winning model was identical for the ‘all’ and ‘even’ conditions but differed slightly for the ‘odd’ trials’ data. However, this difference shows a related family nesting of synaptic groups. To quantify the relative importance of the nested model features, each set can be viewed in isolation in terms of its relative free energy (using all the available data) in Fig. 2C. The schematic Fig. 2D illustrates the layout of these connections, colored according to their relative free energy. Here, superficial AMPA ranks the highest, followed in order by superficial NMDA, deep NMDA, superficial GABA, deep AMPA and finally deep GABA. The ICC was used to assess the reliability of the free energy estimates of the log evidence (Fig. 3A), using ‘odd’ and ‘even’ trials. The reliability was high, in terms of free energy of the full model (with all effects in play), with ICC=0.83 (P<0.0001). We then tested the reliability of the accuracy and complexity that constitute the free energy (where log evidence equals accuracy minus complexity). The accuracy of the states, parameters and the precision and complexity of the states and parameters were again highly reliable (mean ICC=0.85±0.09, P<0.005, Fig. 3B). However, the complexity of precision was not reliable (ICC=0.43, P>0.05).

**Figure 3.**
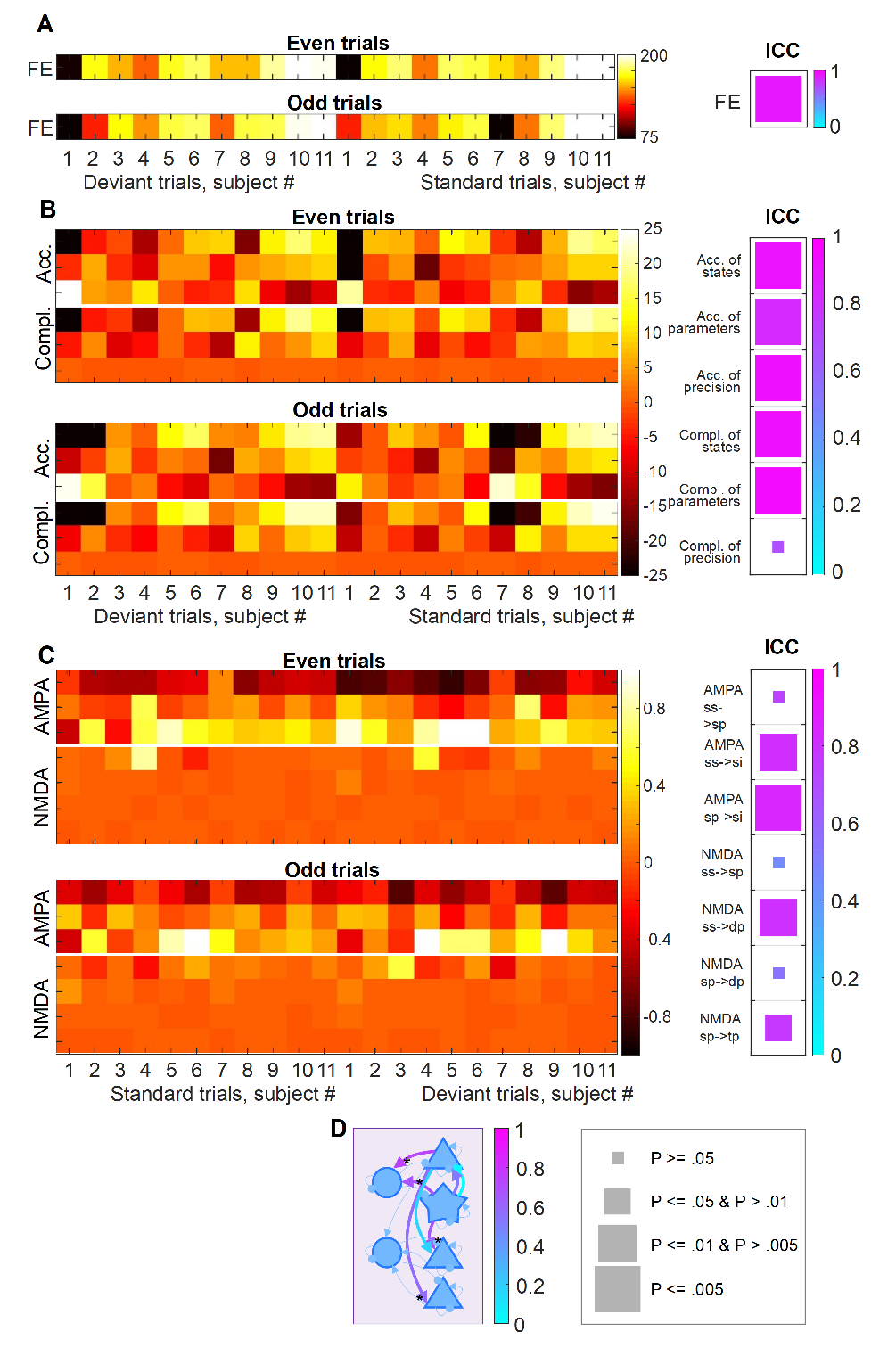
The reliability of free energy, accuracy and complexity estimation. A) The free energy estimates for each subject shown separately by odd and even trials are shown with the ‘hot’ colormap on the left and the ICC reported with the ‘cool’ colormap on the right. For the ICC results, the size of the square relates to its frequentist significance level with the key provided at the bottom of the figure. B) As above for the three accuracy and complexity measures, for the full model. C) As above for the connection set found most likely to be correlated with synaptic density. D) A schematic illustrating the reliable neuronal parameter estimates as thick arrows coloured according to ICC level using the ‘cool’ colormap. *denote the reliable connections.

Having confirmed reliability of model evidence estimates, ICCs were evaluated for DCM synaptic parameter estimates showing the most evidence of regional synaptic density effects (Fig. 3C). Assessing the reliability of single parameter estimates is not the most efficient way to assess reliability, due to conditional dependencies among the parameter estimates. Nonetheless, for 4/7 superficial AMPA, superficial NMDA and deep NMDA connections, the ICC reliabilities were excellent (>0.8). The two superficial AMPA connections and a deep NMDA connection had ICC > 0.6, P<0.005. The schematic in Fig. 3D illustrates the connections with their reliability.

We tested the relationship between the superficial and deep AMPA, NMDA and GABA connections and the two independent measures of disease severity: (1) the Frontal Assessment Battery (FAB) and (ii) the maximal deflection in the ERF during the mismatch window (Ymax). A similar profile was revealed (Fig. 4A), which was highly correlated with the [^11^C]UCB-J result (Fig. 4B, *upper*). Although both independent measures evidenced an effect on synaptic parameters, they did not correlate strongly with each other (Fig. 4B, *lower*).

**Figure 4.**
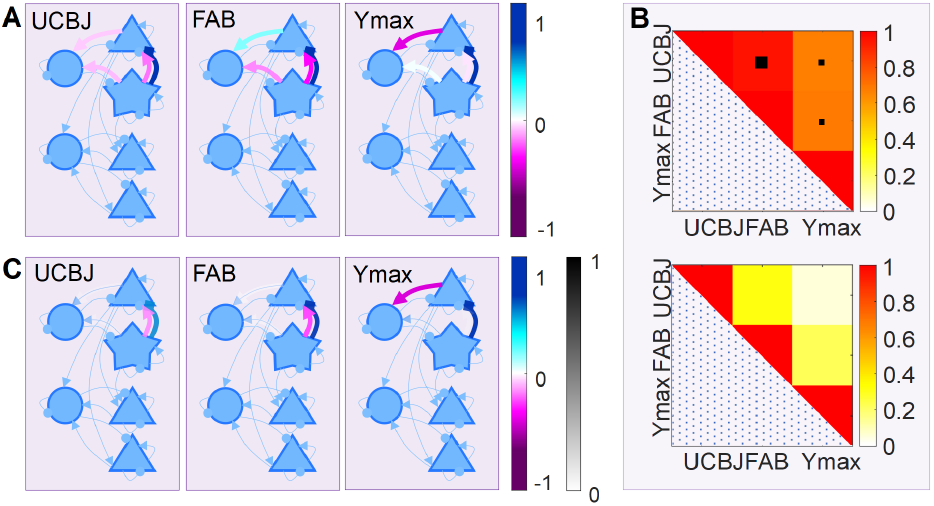
The relationship between connectivity, synaptic loss, clinical impairment and evoked responses. A) The connections found to be significantly correlated with [^11^C]UCB-J, FAB scores and the maximal mismatch-window deflection in the ERF (Ymax) are shown as thick arrows, coloured according to correlation magnitude. B) *Upper*: Correlation matrix created from parameter values for the three covariates. Larger black box denotes P<0.001, smaller black box denotes P<0.05. *Lower*: Correlation matrix for the three covariates. C) Connections that are likely to be related to clinical, synaptic and evoked response metrics following Bayesian model reduction, coloured according to correlation magnitude and transparent according to posterior probability.

Finally, we assessed whether all connections were necessary to explain the above effects of synaptic density and measures of disease severity, or whether some connections could be eliminated as redundant (i.e., increasing model complexity more than accuracy). We used Bayesian Model Reduction (BMR) to find the best PEB model, after removal of redundant connections (Fig. 4C): (i) synaptic density, Ymax and FAB were positively correlated with NMDA activation of superficial pyramidal cells by projections from layer 4 stellate cells; (ii) synaptic density and FAB were negatively correlated with AMPA activation of superficial pyramidal cells by projections from layer 4 stellate cells; and (iii) Ymax was negatively correlated with AMPA activation of superficial interneurons by projections from superficial pyramidal cells.

## Discussion

There are three principal findings of this study—based on the integration of PET measures of neuropathology with MEG measures of pathophysiology— using PEB-DCM^33^. First, regional synaptic density had an effect on the functional synaptic gain in a neurotransmitter- and laminar-specific fashion. Specifically, the superficial and granular glutamatergic synaptic efficacy in the intrinsic connections of an inferior frontal gyrus source—inferred from DCM—was a function of the local synaptic density as measured by [^11^C]UCB-J PET. This corroborates the region- and laminar-specific *post mortem* findings in frontotemporal lobar degeneration^16^, where both AMPA and NMDA receptors are reduced in frontal lobes^34,35^. Second, we found that even though synaptic density, cognition and MMN responses were not strongly correlated with each other, their effects were mediated by very similar local synaptic gains. Third, the DCM approach was highly reliable in terms of estimating model evidence, which is necessary to test hypotheses through model selection (and model reduction). The DCM was highly reliable (for the full model), in terms of the accuracies for the parameters, precision and states. Even at the level of some individual synaptic connections (e.g., AMPA and NMDA), reliability can be high despite the posterior dependencies and multivariate context in which these parameters were estimated.

The relationship between synaptic density and functional change has been examined previously through correlational methods. For example, in Alzheimer’s disease^6,36^, progressive supranuclear palsy^10^, and frontotemporal dementia^12^ synaptic density correlates with cognitive function. Magnetoencephalographic evidence of abnormal oscillatory dynamics has been linked to lower [11C]UCB-J uptake in the occipital cortex^7^, while tauopathies have been correlated with spectral differences and spectrally-constrained changes in connectivity in a range of neurodegenerative disorders; including Alzheimer’s disease and frontotemporal dementia^21,37–41^. These correlative approaches however do not directly support inferences on the pathophysiological mechanisms. In preclinical transgenic tauopathy models, it has been possible to study the mechanisms of abnormal neuronal dynamics, confirming the neurophysiological consequences of pyramidal cell depletion and their reduced synaptic density^42–44^. Despite the presence of tauopathy, these models differ from sporadic human PSP. We chose PSP as an exemplar condition because of the high clinicopathological correlations and mildness of cortical cell loss, despite marked neurophysiological and cognitive changes associated with the prefrontal cortex^25,45,46^.

PSP impairs cognition, particularly in domains associated with frontal cortex such as executive function, cognitive flexibility, response inhibition, verbal fluency and social cognition^22,25,47–49^. These deficits are common in PSP Richardson’s syndrome, and prominent at the presentation of PSP-Frontal phenotype (but not restricted to it). The severity of cognitive change, despite the generally mild cortical atrophy, led to the hypothesis that the impact of PSP on cognition and cognitive physiology is the result of cortical synaptic loss^10^. By translating the synaptic loss in PSP into a generative model of cortical neurophysiology, one can begin to focus on candidate solutions with targeted pharmacology^19^, and link to preclinical models of the synaptopathy in genetic tauopathies^50^. The formal integration of synaptic density into the canonical microcircuit, in which synaptic density forms subject- and cell-specific empirical priors on the microcircuit, goes beyond previous correlative approaches. Given the multivariate nature of the cortical circuits, the use of model comparison—rather than univariate analyses of mean *a posteriori* parameter estimates— properly accommodates the posterior covariance among parameters and increases reliability; two highly desirable properties when anticipating interventional studies.

We probed the cortical circuits using responses evoked in the roving auditory mismatch paradigm. Such tasks and change detection paradigms have been used to study many forms of dementia, ageing and other neurological diseases^51–54^. This paradigm reliably evokes signals in temporal, parietal and frontal regions^55^. The relative simplicity of the task and robustness of the activity it generates makes the paradigm highly suited to these types of modelling^19,31^.

Due to the canonical nature of the local network and the optimization procedure for inversion between the model and MEG (or EEG) data, there is considerable potential for the extension of this method to other disorders, other brain regions, and other multi-modal markers of pathology. The pathology-enriched dynamic causal modelling approach could include other subject-specific markers of pathology, such as magnetic resonance spectroscopy estimates of principal neurotransmitters^46^, or PET ligand markers for the severity of amyloid or Tau burden^56^ or neuroinflammation^57^; or even post-mortem quantitative pathology^58^. Other anatomical regions may be more relevant to hypotheses or mechanisms of other diseases, but the DCM method can be applied to other single regions, or a set of regions connected in networks of distributed sources; with the caveat that increasing complexity of the requisite DCMs may reduce reliability of model inversion.

There are limitations to the study. It is based on data from a small cohort, which could raise the question of type II power for frequentist tests. However, our conclusions are not based on frequentist statistics or the rejection of a null hypothesis. Rather, the ‘power’ in the Bayesian approaches used here derives from making the best sense of data, by posing constrained and informed questions in the form of models or hypotheses and comparing the resulting model evidence. The variational free-energy (i.e., a lower bound approximation to log model evidence) differences reported above means that there is sufficient evidence for hypothesis testing in this cohort, even when accounting for the random effects of being a particular subject, implicit in the PEB analyses. This is not a surprise in view of the severity of PSP, and large effect sizes for both synaptic loss and neurophysiological change^10^. A related issue is the reliability of the DCM approach, which is not guaranteed in this sort of complex system of modelling. However, we provide evidence of excellent reliability, in terms of the inferences based on model selection and high reliability of many individual parameters.

A second limitation is that diagnosis was based in clinical criteria, without neuropathology in most of our cases. However, the cases are typical of PSP, which has the very high clinicopathological correlation expected of Richardson’s syndrome, noting that the cognitive changes are common in those with Richardson’s syndrome and not confined to those with PSP-Frontal phenotype^59^. Third, there is the potential for off-target binding with PET ligands. However, [^11^C]UCB-J has been shown to be reliable and highly correlated with other synaptic markers like synaptophysin^60^, and none of the participants were taking a drug treatment known to interfere with the binding of [^11^C]UCB-J (such as levetiracetam). There are also limitations of the neuronal model. We used canonical microcircuit models, extended in accordance with the favourable model-evidence in Adams *et al*.^31^. However, these neural mass models are approximations and aggregate many cells and cell types within the broad categories set out. Other connections and neuromodulators may exist, for the task and brain regions concerned. Despite their simplification models are useful to characterise features of a system under investigation. Our biophysically informed and constrained model aims to recapitulate the neuronal dynamics of the MMN paradigm, with statistical economy, but we recognise that hypotheses related to other hidden dynamics may call for modification of the model, or analysis of other cortical regions.

In summary, the current study highlights the potential of pathologically-enriched dynamic causal models to elucidate the mechanisms of human neurodegenerative disease. The methodology is suitable for use with other biomarkers of pathology, from PET or spectroscopy, and the assessment of selective pharmacological interventions that target the mechanisms installed in the model. As a potential platform for experimental medicine, the methodology shows good reliability within session, but future assessment of reliability between sessions and during longitudinal follow-up will be useful. We suggest that this methodology could assist the assessment of novel therapies emerging from pre-clinical disease models, aiming to preserve or restore human cognitive neurophysiology.

## Data Availability

The DCM used in this study was adapted from https://gitlab.com/tallie/edcm, as described in Adams et al.30 with prior model parameters altered according to Supplementary Table 2.

## Acknowledgements

We thank the PSP Association & FTD Support Group for raising awareness of the study.

## Supplementary Materials

Supplementary tables are given in the attached document.

---

**Supplementary Table 1. Demographics and test scores**.

Averaged demographic and cognitive data for participants. The mean is shown followed by the standard deviation in brackets.

**Supplementary Table 2. Initial model parameters**.

The initial mean values of notable model parameters and their variance.

