## Supplementary material for "Neurophysiological consequences of synapse loss in progressive supranuclear palsy": SupplTables

**Supplementary Table 1: Demographics and test scores**

| M:F | 6:5 |
| --- | --- |
| Age | 67.5 (10.2) |
| Education (years) | 11.2 (1.5) |
| ACE-R Total (max=100) | 85.6 (6.5) |
| MMSE (max=30) | 28.1 (1.4) |
| Attention (max=18) | 17.6 (0.5) |
| Memory (max=26) | 22.5 (3.6) |
| Fluency (max=14) | 6.5 (3.0) |
| Language (max=26) | 24.2 (1.3) |
| Visuospatial (max=16) | 14.8 (1.2) |
| FAB (max=18) | 14.1 (2.5) |
| INECO Total (max=30) | 19.9 (2.5) |
| Graded Naming: # correct | 19.8 (3.8) |
| Graded Naming: # errors | 10.2 (3.8) |
| PSPRS (max=100) | 24.6 (10.0) |
| Hayling Total scaled scores | 14.8 (3.4) |
| Hayling overall scaled score | 4.4 (1.7) |
| CBI Total (max=180) | 40.4 (25.7) |
| FRS Percentage | 55.1 (29.1) |

**Supplementary Table 2: Initial model parameters.**

| **Parameter grouping** | **Specific parameter** | **Default value** | **Prior Variance** |
| --- | --- | --- | --- |
| Hyperparameter | Precision of the noise | 4 | 1/100 |
| Cell influence on dipole | Stellate (J) | 0.2 | 1/45 |
|  | Superficial pyramidal (J) | 0.8 | 1/45 |
|  | Superficial interneuron(J) | 0 | 1/80 |
|  | Deep pyramidal (J) | 0.2 | 1/45 |
|  | Deep interneuron (J) | 0 | 1/80 |
|  | Thalamic projection (J) | 0.2 | 1/45 |
|  | Leadfields | 1 | 1/45 |
| Network connection gains | Intrinsic gains | 1 | 1/8 |
|  | Extrinsic gains | 1 | 1/8 |
| Decay constants, τ (ms) | AMPA τ | 4 | 0 |
|  | NMDA τ | 100 | 0 |
|  | GABAA τ | 16 | 0 |
|  | GABAB τ | 200 | 0 |
|  | M-current τ | 160 | 0 |
|  | H-current τ | 100 | 0 |
| Miscellaneous strengths | K + leak G | 1 | 0 |
|  | Background V | 2.17 | 0 |
| Reversal potentials (mV) | Na2 + reversal | 60 | 0 |
|  | Ca2 + reversal | 10 | 0 |
|  | Cl reversal | −90 | 0 |
|  | K + reversal | −70 | 0 |
|  | IH reversal | −100 | 0 |
| Firing threshold (mV) | VT (all pops) | −40 | 0 |
| Firing precision | VX (all pops) | 1 | 0 |
| IHI-V slope | VHX | 300 | 0 |
| Cell Capacitances (pF) | Stellate (C) | 200 | 0 |
|  | Superficial pyramidal (C) | 150 | 0 |
|  | Superficial interneuron(C) | 50 | 0 |
|  | Deep pyramidal (C) | 400 | 0 |
|  | Deep interneuron (C) | 50 | 0 |
|  | Thalamic projection (C) | 200 | 0 |
| Delays (ms) | Intrinsic | 2 | 0 |
|  | Extrinsic cortico-cortical | 16 | 0 |
|  | Extrinsic thalamo-cortical | 80 | 0 |
